# MODELLING PRESYMPTOMATIC INFECTIOUSNESS IN COVID-19

**DOI:** 10.1101/2020.11.01.20224014

**Authors:** Russell Cheng, Christopher Dye, John Dagpunar, Brian Williams

## Abstract

This paper considers SEPIR, the extension of an existing parametric SEIR continuous simulation compartment model. Both models can be fitted to real data as they include parameters that can simply be estimated from the data. However SEPIR deploys an additional presymptomatic (also called asymptomatic) infectious stage that is not included in SEIR but which is known to exist in COVID-19. This stage is also parametrised and so can be fitted to data. Both SEPIR and the existing SEIR model assume a homogeneous mixing population, an idealisation that is unrealistic in practice when dynamically varying control strategies are deployed against virus. This means that if either model is to represent more than just a single period in the behaviour of the epidemic, then the parameters of the model will have to be time dependent. This issue is also discussed in this paper.

## 1 INTRODUCTION

A parametric SEIR model has been used by the authors in Dye et al. (2020) to compare the first wave of the COVID-19 epidemics in different European countries. In Dye et al. (2020) this model is fitted to data using the method of maximum likelihood estimation rather than perhaps the more popular Bayesian Markov-chain. The compartmental structure of the SEIR model is standard which means that it does not include a specific compartment to represent the presymptomatic (also asymptomatic) infectiousness stage known to occur in those infected by COVID-19. We describe the SEIR and SEPIR models in Sections 2 and 3, focusing on the models themselves rather than on the effect of the epidemics on the countries it has affected. We discuss the fitting of these models to data, focusing on use of the maximum-likelihood method of estimation which produces (point) estimation of parameter values, as this gives an unequivocal specific model representation of the epidemic. In Section 4 we give a numerical example based on the first wave stage of the COVID-19 epidemic in Switzerland.

An important aspect of the basic maximum-likelihood method is that the parameters values are assumed not only to be unknown but to be fixed in value. Similarly, in the Bayesian case, the distributions of the parameters are not only unknown, but are assumed to be fixed. However different strategies varying over time have to be deployed in trying to contain in a fast moving epidemic like that produced by COVID-19. This means that the model parameters do not remain constant, so but have to be time dependent if the trajectory of the epidemic is to be correctly reproduced. Note that use of the models based on SEIR can be used in examining more than one wave, see for example Dagpunar (2020).

For reasons of space and simplicity we discuss in this paper only ‘first wave’ model behaviour, leaving for elsewhere discussion of situations where time-varying parameters might be used.

We do however discuss how progress of an epidemic is summarised by the *effective* reproduction number *R*_*t*_, a dynamically varying version of *R*_0_, the (basic) reproduction number. Theoretically, *R*_0_ is unequivocally defined in terms of the idealized epidemic infecting a population in a homogeneously mixing manner. However when monitoring the progress of an epidemic *R*_*t*_ is more useful, and in lay terms seems to be what is called the reproduction number. It can still be defined to be the expected number of persons infected by an infected individually, but should be time dependent because of changes in the management of the epidemic and in the susceptible population. The calculation should thus hold whether the infection is homogeneously mixing or not. We consider this in Section 5

## 2 THE SEIR MODEL

The SEIR model has been described in the Supplementary Materials of Dye et al. (2020), but for ease of comparison with SEPIR model we give the description again here. The model is of a homogeneously mixing population with four compartments representing those who are susceptible, exposed, infectious and recovered or died (SEIR), as shown in Figure 1.

**Figure 1.**
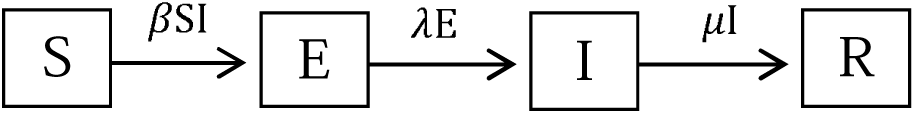
The SEIR model. The compartments denote those in the population that are Susceptible, Exposed, Infected and Recovered.

The variables S, E, I and R satisfy the ordinary differential equations:

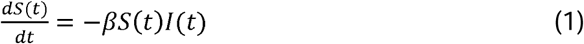

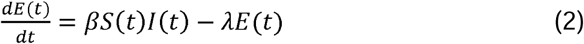

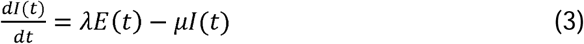

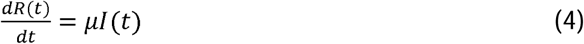

A convenient recent reference is Ma (*3*) who uses a slightly different notation. Also, to highlight deaths due to the virus we divide those that recover well and those that die due to the virus. Thus the infectious are divided into two compartments as illustrated Figure 2.

**Figure 2.**
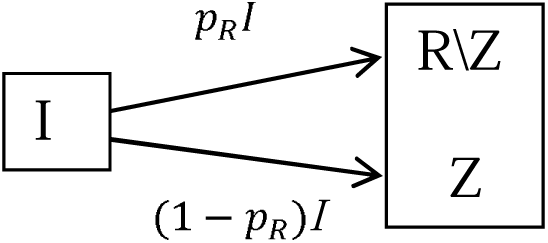
Adjustment of the SEIR model where R is divided into two compartments, R | Z, those that recover and Z, those that die; where p_R_ is the proportion that recover.

More elaborate models can and have been developed. For example, see Dagpunar (*4*) who extends *R* into additional compartments representing different outcomes of hospitalization

The SEIR model of Figures 1 and 2 are assumed to depend on certain parameters, initially assumed unknown. Fitting the model to data, is simply the process of estimating the parameters, either directly using data obtained from observing the epidemic, or from information obtained from other sources. Once the parameter values are estimated, the behaviour of the SEIR model is completely specified. The parameters are defined in Dye et al. (2020). To avoid repetition and avoid confusion they are not discussed directly here as we shall be discussing the SEPIR model where a very similar set of parameters will be fully defined.

However we do point out here the time-delay parameter *τ* used to modify equation (4) to:

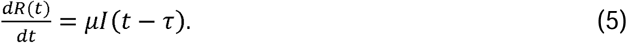

We denote by **θ** = (*b*1, *b*2, …, *b*_*m*_), the vector of parameters, where *m* is the number of parameters. In Dye et al. (2020), *m* = 9. In the SEPIR model of Section 3, *m* = 11. With **θ** given, the four differential equations (1), (2), (3) and (5) can be solved by numerical integration to give the trajectories

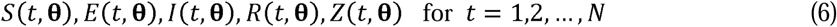

where *t* is the day and *N* is the number of days of interest. We used the standard method of Maximum Likelihood (ML), as given for example in Cheng (2017), to estimate parameter values.

Here we outline the approach used to estimate the parameters from a sample of observed daily deaths, say. Let the sample of observed number of daily deaths be denoted by

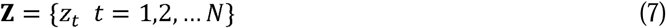

where *z*_*t*_ is the number of deaths on day *t* and *N* is the number of days observed. If the observations were made without error and if, with the right parameter values are correct for **θ**, then the death trajectory {*Z*(*t*, **θ**) *t* = 1,2, …, *N*} would match the observed deaths **Z** in (7). So the model would then be successful in explaining deaths.

To include statistical uncertainty in the model we assume instead

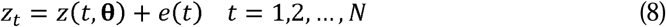

where *e*(*t*) is random error. For simplicity the *e*(*t*) are assumed to be normally and independently distributed (NID) with standard deviation *σ*, i.e.

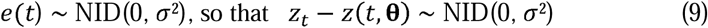

The logarithm of the distribution of the sample is then

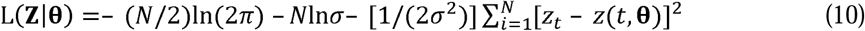

where **Z** is the random argument, and the parameters **θ** are fixed. In ML estimation (MLE), this is turned on its head so that **Z** is simply the known sample of observations now regarded as fixed and we write L as L(**Z**|**θ**) = L(**θ**|**Z**)) calling it the (log)likelihood to indicate that it is now treated as a function of **θ**. The ML estimator 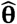 is simply the value of **θ** at which L(**θ**| **Z**) is maximized. i.e.

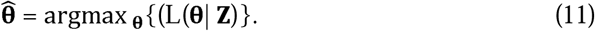

Nelder-Mead numerical search for the maximum was used. This goes through different **θ**_*i*_ *i*=1, 2, 3,… comparing the different L(**θ**_*i*_, |**Z**) to find 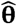, the best **θ**.

To simplify description of the estimation process, only fitting to deaths data, **Z** as in (7) has been described, but the method extends straightforwardly to include other data samples. For example

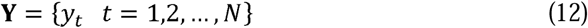

where *y*_*t*_ is the number of active cases on day *t*. Fitting simultaneously to both **Y** and **Z** can be carried out by adding to the right-hand side of (10) a corresponding set of terms for **Y**.

Numerical solution of the differential equations requires initial values for *S, E, I, R*. These are essentially scale invariant with (*S +E +I + R*) constant and independent of *t*. So the numerical integration can conveniently be done using S(0, **θ**) = 1, E(0, **θ**) some small quantity subsequently adjustable as its initial value *e*_0_ is a parameter; with *I* and *R* initially zero. The size of the exposed population, parameter *s*_*0*_, is only needed to provide scaled values *S, E, I, R* at each step for comparison with the data **Y** and **Z**.

## 3 SEPIR MODEL

### 3.1 Structure of the SEPIR Model

In the SEPIR model we introduce an extra compartment to the SEIR model in Fig. 1 changing it to Fig 3:

**Figure 3.**
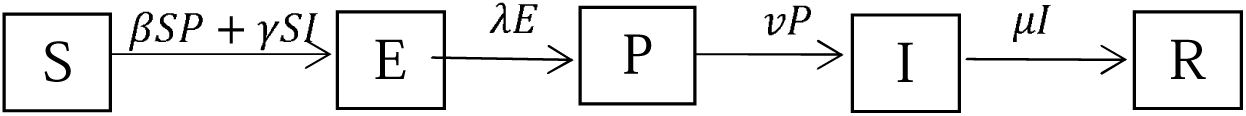
The SEPIR model. The compartment P denotes those who are infectious but are presymptomatic whilst I denotes that are infectious and symptomatic.

The original *I* compartment is now split into two with its first compartment, *P*, comprising those infectious who are pre, i.e. asymptomatic, and the second, *I*, comprising those infectious who display symptoms. The ordinary differential equations (1), (2) and (3) in the SEIR model are replaced by the differential equations (13), (14), (15) and (16), with (4) and (5) remaining unchanged. There are two terms in going from S to E: comprising those infected by someone in P with transmission rate *β*, and those infected by someone in I with transmission rate *γ*. The reciprocal *λ*^−1^ is the mean period someone spends in state (compartment) P whilst *µ*^−1^ is the mean period spent in I.

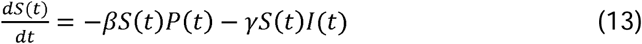

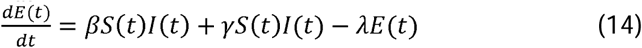

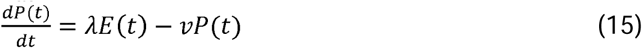

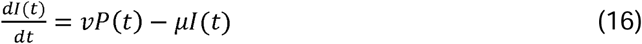

We treat the quantities *β, γ, λ, ν, µ* as parameters to be estimated. However we include six further parameters *t*_0_, *e*_0_, *s*_0_, *σ, p*_*R*_ and *τ*. These are all listed and defined in columns 1 and 2 of Table 1.

**Table 1.** Parameters of the SEPIR model with estimates for Switzerland.

| Symbol | Definition | Estimated value and 95% confidence interval |
| --- | --- | --- |
| $\beta$ | Presymptomatic transmission rate | 0.44 (0.435– 0.454) |
| $\gamma$ | Symptomatic transmission rate | 0.137 (0.128– 0.147) |
| $\lambda^{-1}$ | mean latent period in compartment E | 1.19 (1.13– 1.26) |
| $\nu^{-1}$ | mean presymptomatic period | 4.40 (4.00– 4.62) |
| $\mu^{-1}$ | mean symptomatic period | 14.0 (13.8– 14.3) days |
| $t_0$ | number of days from start of epidemic before observations began | 30 days (too small to measure) |
| $e_0$ | initial number of individuals exposed | 6.6 (5.6– 8.2) E-07 |
| $s_0$ | initial size of exposed population (to scale the epidemic) | 3.67 (3.60 – 3.74) E+04 |
| $\sigma$ | standard deviation of observational error | 103 (82 – 106) |
| $p_R$ | probability of someone infected recovering well | 0.943 (0.942 – 0.945) |
| $\tau$ | mean time between the end of infectiousness and recovering well or death | 3.0 (2.8 – 4.0) days |

There is flexibility in fitting of the model to data. Some of the parameters can be given fixed predetermined values with the others fitted to data by Maximum Likelihood as described in the SEIR model is Section2.

### 3.2 Switzerland: A Numerical Example

Column 3 gives the parameter values when SEPIR was fitted estimating all 11 parameters by maximum likelihood using *N* = 109 days of data based on daily observations starting on 15 Feb 2020. Two series: Daily New Cases and Daily Deaths were used. The values of all the parameters are of interest. The parameter values for Switzerland are given in Table 1. We highlight two aspects.

Firstly consider the population size. In a standard the SEIR because the differential equations are scale invariant, one can assume a notional standard population size of 1. However in our model we allow the population size to be variable with a size estimated by allowing rescaling to maximize the likelihood. The estimated population size of 36,700 is remarkably small suggesting that can be interpreted as the size of the homogeneously mixing population actually ‘seen’ by the virus compared with the actual population size of 8.2 million. Moreover without examining unavailable regional records it may be that the outbreak in Switzerland was mainly confined to parts nearest Italy the latter being the first European country to be badly affected by COVID.

Secondly we examine whether the SEPIR model gives any indication of the extent of the presymptomatic stage.

First we summarize what is already known about this stage by reporting the findings of He at al. (2020) who investigated the case histories of 77 infector-infectee pairs in each of which an infectious person, the infector, goes on to infect a susceptible person, the infectee.

Citing the mean incubation period as 5.2 days, He et al, (2020) estimate the serial interval to be 5.8 days. From this they infer that infectiousness starts 2.3 days after the onset of infection and peaks just 0.7 days before symptom onset, giving an estimated proportion of infections of 44% as occurring before the onset of infector symptoms. Infectiousness then declined within 7 days. Figure 4 is a schematic showing the infector-infectee relationship.

**Figure 4:**
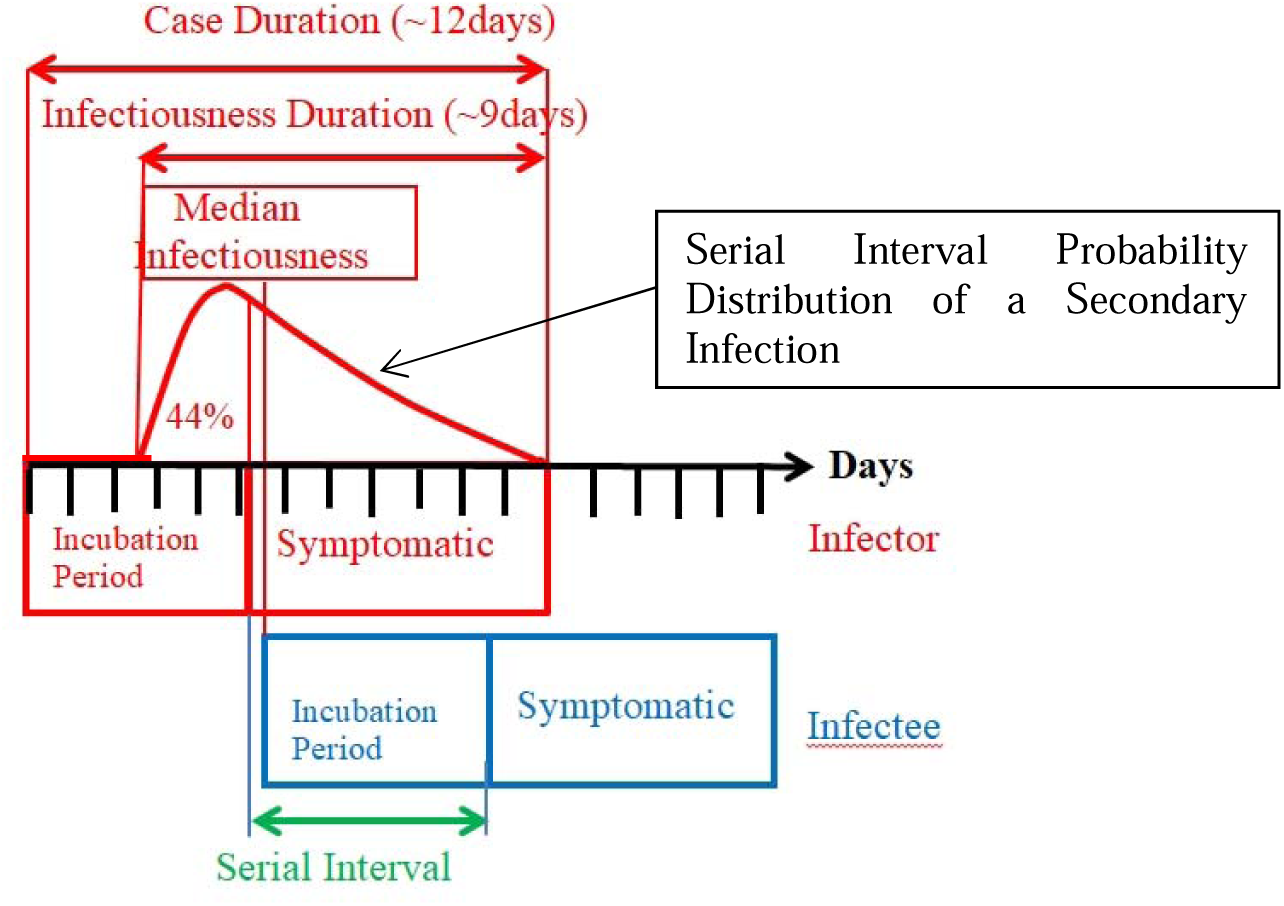
Infector-Infectee Relationship as described by He et al (2020).

The estimate of He et al. (2020) that the proportion of individuals infected presymptomatically is 44% means, in our case, that the proportion (*βν*^−1^)/(*βν*^−1^ + *γµ*^−1^) should therefore be this value at least approximately. From Table 1, the value is 50.4%.

This is very much in accord with the higher presymptomatic infection proportions given by Tapiwa et al (2020): 48% in Singapore and 62% in Tianjin.

The practical consequences of this finding is evident, with elaborate track and tracing required to identify presymptomatic infections.

The quality of the fit achieved by the SEPIR model is illustrated in Figure 5 where the Active Cases and Cumulative Deaths curves obtained by fitting the model to both data sets simultaneously are plotted with their corresponding data.

**Figure 5:**
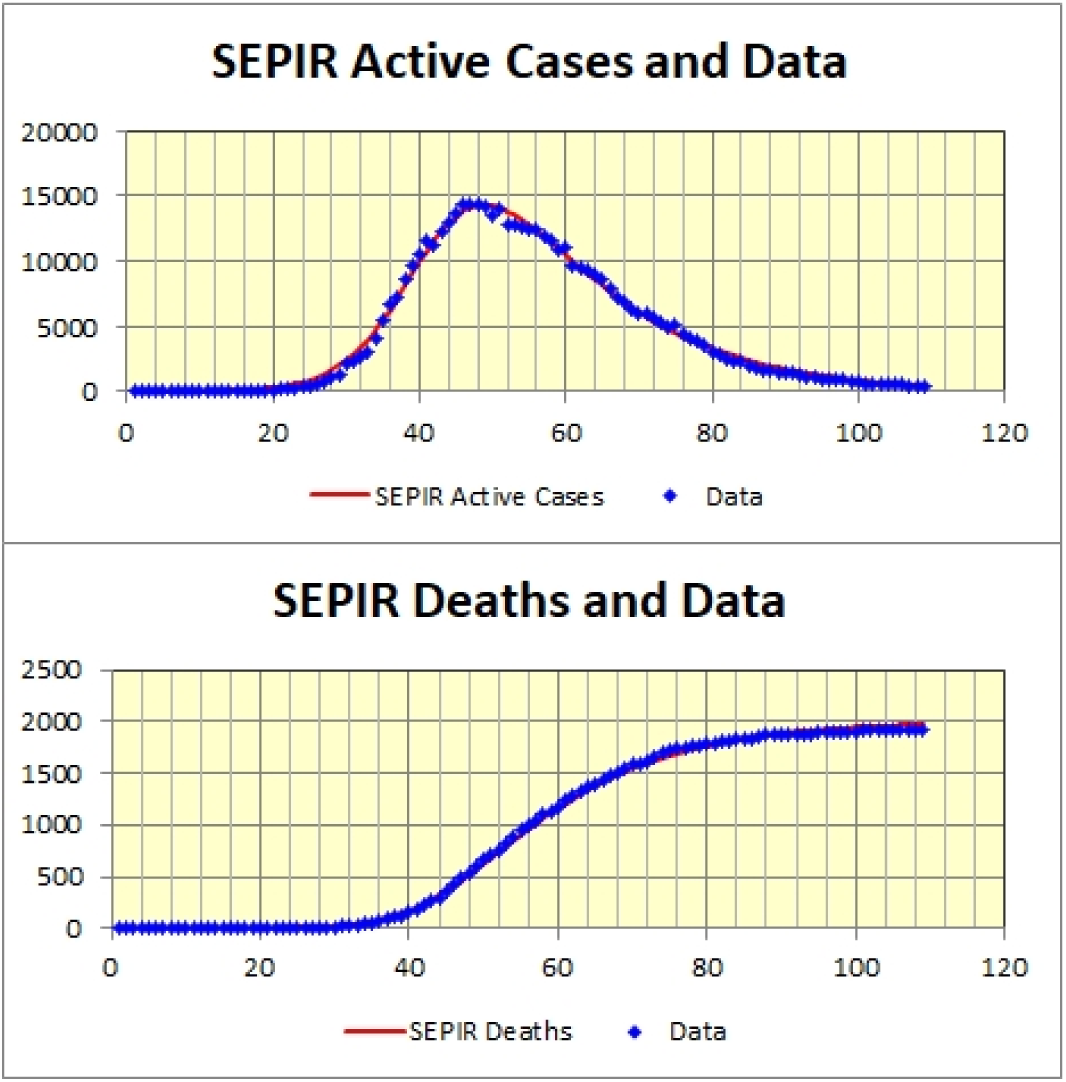
SEPIR Active Cases and Cumulative Deaths fitted to Swiss Data. Horizontal axis is days with day #1 = 15^th^ February 2020.

The SEPIR model was then repeatedly fitted to independent parametric bootstrap replications of the actual observed active cases and cumulative deaths data. As described in Subsection 4.1.3 of Cheng (2017) confidence intervals for the parameters can be obtained from the bootstrap parameter estimates. For illustration, the resulting 95% confidence intervals for each of the fitted parameter estimates are reported in Column 3 of Table 1, using 500 bootstraps.

Charts of fitted SEPIR trajectories provide an easily understood way to display results. For example the fitted SEPIR cumulative deaths trajectory (red) is displayed in Figure 6 together with the observations (black). The method described in Section 4.3 of Cheng (2017) can be used to provide a confidence band for any model trajectory. For example we have a bootstrap cumulative deaths trajectory corresponding to each bootstrap sample. These are plotted (in grey) in Figure 6 giving a bundle of trajectories, with 95% confidence limits (green and blue). Only 250 bootstrap are depicted.

**Figure 6:**
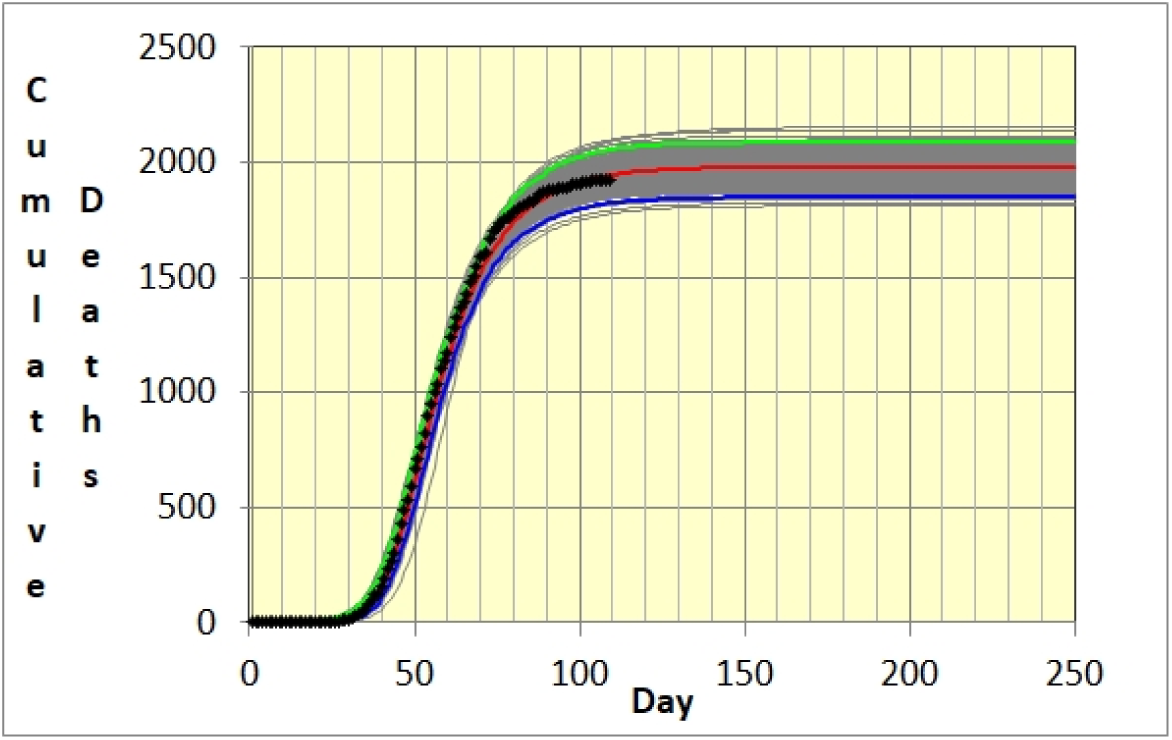
SEPIR Fitted Cumulative deaths trajectory (red) for Swiss data obtained from 109 observations (black). Upper (green) and lower (blue) confidence limits are also depicted.

## 4 *R*_*T*_ THE EFFECTIVE REPRODUCTION NUMBER

In the SEIR model, the Reproduction Number *R*_0_ is simply, but precisely, defined as the number of susceptible individuals that an infectious person will go onto infect when the epidemic first starts, assuming that the population is homogeneously mixing. In the SEIR model *R*_0_ can be obtained, see for example Dagpunar (2020), from the biological transmission rate of the virus β and mean period of infectiousness *μ*^−1^ as

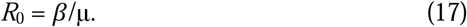

As mentioned in the Introduction, in practice *R*_*t*_, the effective reproduction number, is more useful as, throughout the epidemic, it can be continually used to gauge how well control strategies are working. The theoretical basis underlying the calculation *R*_*t*_ is well described by Ma (2020). We have

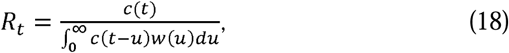

where *c*(*t*) is the incidence curve of new cases at time *t* and *w*(*u*) is the *serial interval probability distribution* of a secondary infection; so that *w*(*u*)*du* is the probability that an infectious individual (the infector)) infects someone else (the infectee) in the time period (*u, u+du*). This probability distribution is depicted in Figure 4.

The denominator in Equation (18) measures how the new cases at time *t* arise from those infected prior to time *t*. The epidemic clearly is rising or falling depending on whether the numerator is larger or small than the denominator, with equilibrium when they are equal. Thus *R*_*t*_ has the critical reproduction property of *R*_0_ but moreover is dynamic, so that it can be used to gauge the progress of the epidemic as this develops.

It turns out that the formula (18) is quite robust so that the serial interval distribution does not have to be estimated all that accurately. In fact Germany, early during its first COVID-19 epidemic wave, used the simple denominator *c*(*t*-4). Cori et al. (2013), though using a Bayesian approach, examined various empirically obtained serial interval distributions drawn from different epidemics. In Dye et al. (2020) the authors used a discretized and shifted gamma distribution *g(t), t* = 1,2, …, 12 to represent the serial interval distribution *w*(*u*) that is shown as the red curve in Figure 4, calculating the denominator as

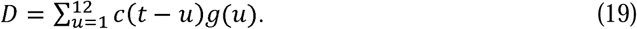

Figure 7 depicts *R*_*t*_ calculated using this formula for Switzerland when c(t) is a daily 7-day moving average of new cases.

**Figure 7:**
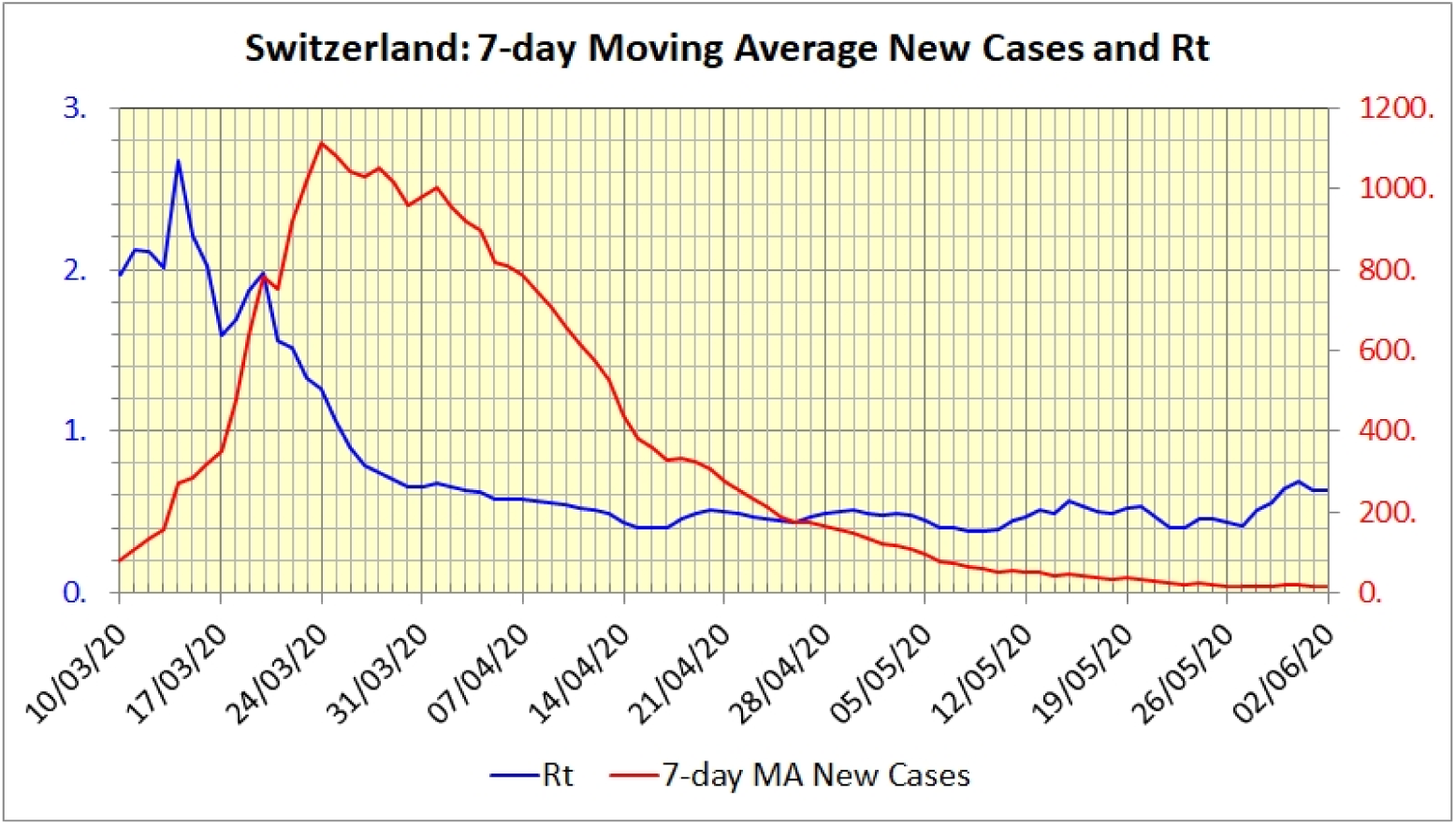
Chart of the effective *R*_*t*_ calculated using the formula in Equation (19) for Switzerland where *c*(*t*) is the 7-day moving average of new cases.

## 5 CLOSING REMARKS

In conclusion the SEPIR model is more flexible than one with parameters both given and fixed. For example, our model estimates an initial susceptible proportion rather than taking it as a given (for example the country’s actual population). However on the other hand our transmission rates, though estimated, are supposed constant rather than time dependent which would be needed to model changing management of the epidemic. These two things could explain why the estimates of some of the biological parameters are rather different from those observed in some other studies.

We end with two warnings.

Regarding just model fitting alone, this is of academic interest in its own right. We have not examined in detail the robustness of maximum likelihood optimization. In our numerical example we chose the first wave of the epidemic in Switzerland because the data corresponded well to the characteristics of the SEPIR model. However even in this example alternative good fits can be achieved with combinations of parameter values different from those reported in Table 1. Thus, in practice, comparison with parameter estimates obtained in other ways should always be made where possible to assess when our estimates can be relied on.

More generally the simplicity of models such as SEIR or SEPIR. means that the practical usefulness of using them on their own, in isolation, is limited. The models are idealizations of the way the epidemic behaves and of population behaviour. This latter is particularly important and will depend on control policies and their timing. Lack of homogeneity of population behaviour is an important factor in implementing control policies because these have to recognize the political issues they give rise to for the population as a whole to be willing to follow them. To make a national control policy fair, a detailed model allowing for regional differences may well be needed.

## Data Availability

The Swiss data, ( 109 days of new cases and deaths starting from 15 Feb 2020) referred to in the article is publically available online.

https://ourworldindata.org/coronavirus-source-data

## AUTHOR BIOGRAPHIES

**RUSSELL CHENG** retired from the University of Southampton in 2007 where he had been Head of the Operational Research Group, having held previous positions at Cardiff University and the University of Kent at Canterbury. https://www.southampton.ac.uk/maths/about/staff/rchc.page

**CHRISTOPHER DYE** FRS, FMedSci, has held positions at the London School of Hygiene and Tropical Medicine, the World Health Organization, Gresham College London. He has been Visiting Professor of Zoology at Oxford since 2009, and became a Visiting Fellow at the Oxford Martin School in 2019. http://en.wikipedia.org/wiki/Christopher_Dye

**JOHN DAGPUNAR** retired from Edinburgh University in 2008. He is Visiting Research Fellow in Mathematical Sciences at the University of Southampton. His research interests are in simulation, financial mathematics, health studies, and reliability. https://www.southampton.ac.uk/maths/about/staff/jd2y15.page

**BRIAN WILLIAMS** is Senior Research Fellow at the South African Centre for Epidemiological Modelling and Analysis (SACEMA) having held the position of Epidemiologist at the World Health Organisation from which he retired in 2008.

## REFERENCES

Cheng R C H (2017). Non-Standard Parametric Statistical Inference. Oxford University Press, Oxford.

Cori A, Ferguson N M, Fraser C and Cauchernez S (2013). A new framework and software to estimate time-varying reproduction numbers during epidemics. American Journal of Epidemiology 178(9): 1505–1512.

Dagpunar J S (2020). Sensitivity of UK Covid-19 deaths to the timing of suppression measures and their relaxation. Infectious Disease Modelling 5: 525–535 https://doi.org/10.1016/j.idm.2020.07.002

Dye C, Cheng R C H, Dagpunar J and Williams B G (2020). The scale and dynamics of COVID-19 epidemics across Europe. medRxiv https://doi.org/10.1101/2020.06.26.20131144.

He L, Lau E H Y, […], Leung G M (2020). Temporal dynamics in viral shedding and transmissibility of COVID-19. Nature Medicine 26: 672–675.

Ma J (2020). Estimating epidemic exponential growth rate and basic reproduction number. Infectious Disease Modelling 5: 129–141.

Tapiwa G, Kremer C, Chen D, Torner A, Faes C, Wallinga J and Hens N (2020). Estimating the gerneration interval for coronavirus disease (COVID-19) based on symptom onset, March 2020. Euro Surveill. 25(17):pii=2000257.

